# Predicting Alzheimer’s Disease Diagnosis, a Decade or more Years before Onset using the Electronic Health Record and Random Forest Machine Learning Models

**DOI:** 10.1101/2025.11.04.25338396

**Authors:** Sanya B. Taneja, Richard D. Boyce, Scott A. Malec, Steven M. Albert, C. Elizabeth Shaaban, Arthur S. Levine, Paul Munro, Jiang Bian, Jie Xu, Demetrius Maraganore, Karen Schliep, Jonathan C. Silverstein, Michelle Kienholz, Helmet T. Karim

## Abstract

**INTRODUCTION:** There is need to detect and intervene in pre-clinical phases of Alzheimer’s disease (AD). Electronic health records (EHRs) may help predict AD using machine learning methods.

**METHODS:** We identified EHRs for 19,473 cases with AD and 111,922 controls. Records spanned 10 or more years prior to AD diagnosis. We trained a random forest model (employing 5-fold cross-validation with 2,499 features) to predict AD 10 years prior to its onset using a 75/25% train/test split and then computed permuted feature importance.

**RESULTS:** We achieved an area under the ROC curve of 0.80. Feature importance identified factors associated with AD, including age, sex, race, ethnicity, BMI, cardiovascular diseases, inflammation, pain, sleep and mood disorders, trauma, other neurodegenerative disorders, diuretics, colon-related disorders and procedures, seizures, and vitamin B12.

**DISCUSSION:** This is the first EHR-based model to predict AD 10 years prior to onset, which could help predict AD and inform prevention/early intervention.

## Background^a^

Alzheimer’s disease (AD) affects 7.2 million people in the United States, growing to an expected 13.8 million in 2060, and is the seventh leading cause of death(1). There are currently only limited treatment options for individuals with AD that have modest effects on cognitive function (2-4). There is a critical need to develop tools to detect AD earlier, identify protective and risk factors of AD, and intervene earlier, ideally prior to the onset of AD pathology. Indeed, sufficient evidence indicates that amyloid positivity, levels of some experimental biomarkers(5-8), and even some clinical signs precede clinical diagnosis of AD by decades (9, 10). The electronic health record (EHR) is a promising source of patient data that, combined with machine learning, can inform, or enable clinical interventions that could help improve the health of patients at high risk of AD. In addition, the EHR may help identify patients who are at-risk for improving study recruitment for clinical trials and studies on high-risk patients.

For example, many clinical trials are currently focusing on amyloid-clearing treatments. These studies have shown adequate clearance of amyloid but only relatively minor improvements in cognitive function. Early detection could also help inform more holistic treatment or prevention strategies, where teams of healthcare workers can work in tandem to treat several risk factors with machine-guided and personalized precision(11). Numerous factors contribute to cognitive impairment and rapid cognitive decline – either as part of the central mechanisms (e.g., affecting amyloid or tau) (12) or other lifestyle and behavioral factors. Fourteen major modifiable risk factors estimated responsible for approximately 45% population risk in high-income countries such as the United States have been identified including low education, hearing loss, LDL cholesterol, depression, traumatic brain injury, physical inactivity, diabetes, smoking, hypertension, obesity, alcohol, social isolation, air pollution, and visual loss (13). The Electronic Health Record (EHR) is highly effective in facilitating predictive modeling, given that it accumulates data in real time over extended periods as an integral component of clinical practice. The nature of EHR data makes it exceptionally compatible with machine learning techniques, enabling researchers to construct models that pinpoint factors associated with AD.

Several studies have focused on predicting AD diagnosis using EHR data(14-16). One study identified 2,640 AD patients and 40,736 records to predict AD at 1, 2, 3, and 4 years using EHR data; these models achieved an area under the receiver operating characteristic curves (AUCs) of 0.68-0.78(15). Another group identified patients with mild cognitive impairment (MCI) using clinical notes and deep learning, achieving an AUC of 0.75(14). Yet another group used graph neural networks to predict AD in a sample of 1.64 million records with 8,174 AD patients and achieved an area under the precision-recall curve (AUPRC) of 0.39-0.71(16).

Two more recent studies have evaluated data up to 5 years in advance. One study used 35,866 cases and 179,330 controls in an administrative claims dataset and predicted AD 4-5 years in advance with an AUC of 0.69(17). Another evaluated the effectiveness of predicting AD diagnosis 5 years prior to onset, which is the furthest back to date(18), using 23,835 cases and 1,038,643 controls, making it one of the largest to date. They achieved high prediction accuracy – AUC of 0.85 for 5 years prior to onset – and identified several predictive factors. An even more recent study identified predictors up to 7 years prior to onset(19) achieving an AUC of 0.72 in individuals with more accurately defined AD diagnosis through a memory and aging center.

In this study, we identified a large set of patient records spanning more than a decade of continuous care for 19,473 cases and 111,922 controls. The goal of the study was to use machine learning to predict AD 10 or more years prior to diagnosis (e.g., a patient with a diagnosis in 2016 had to have EHR data from at least 2006 or earlier).

## Methods

### Extracting Electronic Health Record Data

An EHR dataset was extracted from the UPMC EHR using the Health Record Research Request service(20). The University of Pittsburgh Institutional Review Board approved this study as exempt and confirming that consent was not necessary. Data were de-identified to a HIPAA-limited dataset and contained information on patient demographics, allergies, diagnoses, family history, immunizations, lab results, medications, medical history, procedures, social history on alcohol and tobacco use, and vitals. The data included all types of visits (e.g., inpatient, outpatient, emergency, etc.). We used the following diagnostic codes to identify diagnoses of dementia or AD (International Classification Code of Disease (ICD)-9 and 10 codes 290.0, 290.1, 290.10, 290.11, 290.12, 290.13, 290.20, 290.21, 290.3, 294.20, 294.21, F03.90, F03.91, G30, G30.0, G30.1, G30.8, and G30.9). The case cohort included patients diagnosed with AD or dementia from 1/1/2016 to 12/31/2021 who had at least one UPMC EHR encounter 10 or more years before this diagnosis.

This extraction resulted in 19,473 cases and 111,922 controls matched on the index date—or the date of diagnosis in AD cases. In the final pull of data, controls were excluded for exposure to acetylcholinesterase inhibitor or memantine or any of these diagnostic codes: 331.0, G30, G30.0, G30.1, G30.8, G30.9, F00, F00.0, F00.1, or F00.2 (Alzheimer’s Disease).

### Preprocessing of the EHR data

All EHR data were transformed to the Observational Data Health Sciences and Informatics (OHDSI)’s Observational Medical Outcomes Partnership (OMOP) Common Data Model (CDM) (version 5) with standardized vocabularies for each data domain. The final data were stored in the CDM person (demographics), condition occurrence, drug exposure, measurement, observation, procedure occurrence, visit occurrence, condition era, and drug era tables. The condition era and drug era tables are chronological periods of condition occurrence and drug exposure, respectively. We manually mapped codes for family history concepts, allergies, and encounter types to standardized vocabulary concepts as these were not originally present in the data. We also used term-matching to find standardized concepts for unmapped National Drug Code drugs, which resulted in a standardized representation of the data using this and other vocabularies, such as the Systematized Nomenclature of Medicine, RxNORM, and the Logical Observation Identifiers Names and Codes. It also enabled using the OHDSI Patient-Level Prediction R package for predictive analysis(21).

### Training of Random Forest Machine Learning Model

We trained a random forest model to predict AD diagnosis (cases vs. controls) 10 or more years prior to the AD diagnosis date (observation window). We chose a random forest model as random forests have been shown to outperform several other methods on tabular data including deep nets(22). The data from the start of a person’s EHR history to 10 years prior to diagnosis of AD were collapsed into features for the random forest model. For each measurement, condition, procedure, drug, visit, and observation record in the data, the values in the observation window were converted to binary values representing present/absent in the EHR. The values for each measurement were included along with an indicator as to whether the measurement value was below, within, or above the normal range. We also included as features in the model the age groups, race and ethnicity categories, number of visits, Charlson Comorbidity Index score, Diabetes Complication Severity Index, CHADS_2_, and CHA_2_DS_2_-VASc scores.

We used a 75% training and 25% test split of the data to train the model and evaluate performance, respectively. The training was conducted via 5-fold cross-validation on the training set only. This prediction used 20,931 features. The minimum covariate occurrence had to be above 0.001; covariates that occurred in a fraction of the target population less than this value were removed. This resulted in 2,499 features that were included in the final model. We used the OHDSI Patient-Level Prediction R package to train and test the random forest model(21). The R package is specifically designed to train and validate models for data represented in the OMOP CDM format. We conducted hyper-parameter optimization over the following: number of trees (50, 100, 500, 1000), depth of trees (4, 10, 17, 25, 50), and number of features per tree (5, 20, 100, 200, and square root of the total number of features). The model was optimized using the AUC value.

### Evaluation of Random Forest Model

We evaluated the random forest model over the test set using AUC and AUPRC, accuracy (# predicted accurately/total), positive predictive value (PPV, # AD predicted accurately/total # predicted AD), negative predictive value (NPV, # controls predicted accurately/total # predicted controls), sensitivity (# AD predicted accurately/total AD), and specificity (# controls predicted accurately/total controls). We computed the optimal threshold obtained from the training data and evaluated the above metrics on the test data. The optimal threshold was obtained by maximizing the Youden Index, the threshold optimizing model performance when equal weight is given to sensitivity and specificity. The test set was only used to evaluate these after completing all training.

### Ranking Feature Importance

We conducted a permutation feature importance (PFI) analysis on both train and test data. PFI permutes each feature, computes the AUC, and then compares the result with the original model’s AUC to the permuted model’s AUC. The PFI for each feature is then calculated as the original AUC minus permuted AUC. Larger reductions in AUC indicate a more important feature. To identify which features to focus on, we sorted the PFI (descending) and then walked along the cumulative sum curve of the PFI one bisection point at a time. At each point, two lines are fit: one to all the points on the left and one to the right. The knee is the point where this bisection minimizes the sum of errors for the two fits. This approach identified the inflection point of the PFI curve. We included only those features that were ‘above’ this inflection point for interpretation.

## Results

The characteristics of AD cases and controls are shown in Table 1. The cases and controls are predominantly non-Hispanic white, with >60% females and a large age distribution.

The performance of the random forest model on the training data was AUC of 0.85 and AUPRC of 0.68. On the test set, performance was AUROC 0.80 and AUPRC of 0.55. The optimal prediction threshold on the test dataset was 0.25 which maximized the balanced F measure at a value of 0.47. At this threshold, the model achieved a PPV of 0.53, NPV of 0.90, sensitivity of 0.43, and specificity of 0.93 (see Figure 1). The optimal random forest model used for prediction included 1000 trees, a maximum depth of 25, and a number of features equal to 200 after hyperparameter optimization.

**Figure 1.**
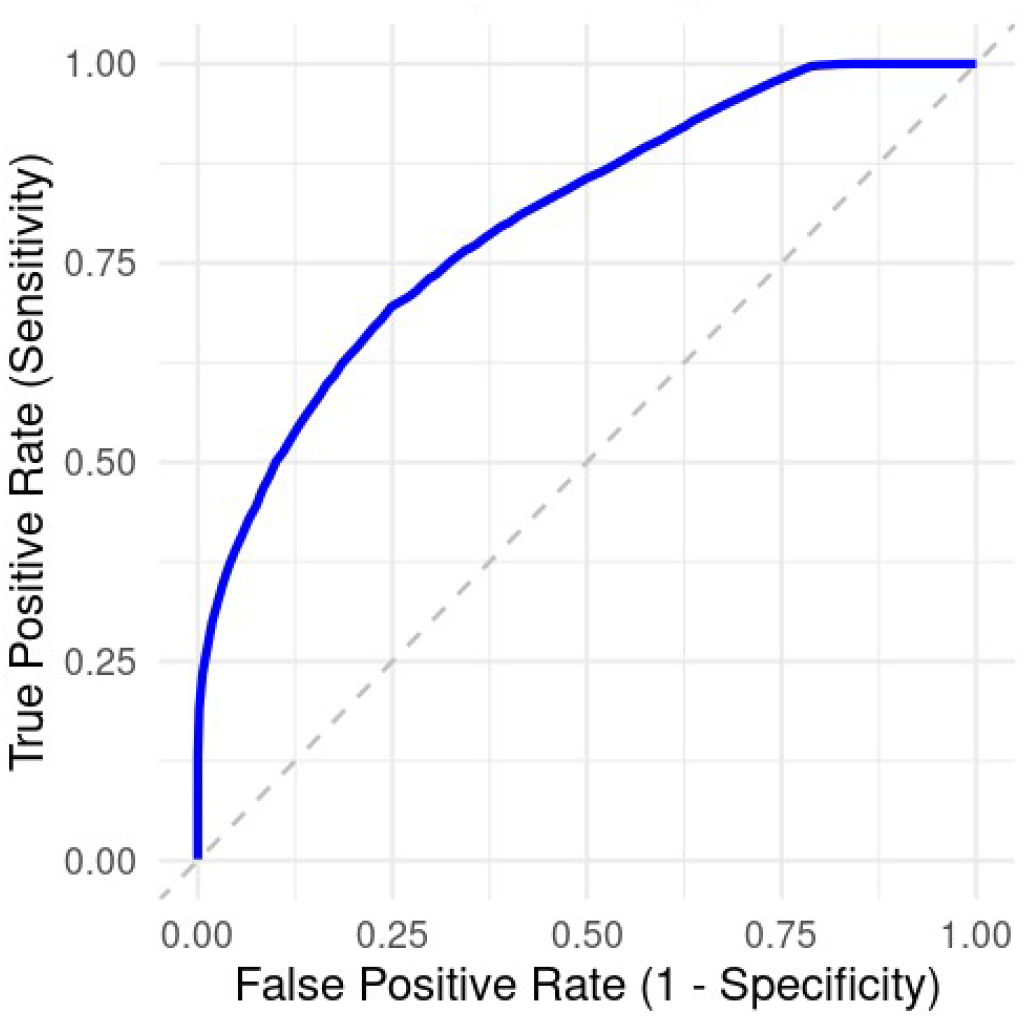
Area under the ROC curve for the AD random forest machine learning model on the test set plotting the false positive to true positive rate at various thresholds. The threshold that optimized the balanced F-measure was 0.25 which led to an F-measure of 0.47, sensitivity of 0.43, specificity of 0.93, PPV of 0.52, and NPV of 0.90. Dotted line represents chance prediction.

Using the knee point method, we found that the PFI of the top 227 features was most important. We report these in Table 2. Broadly, this included a number of categories, which we summarize:

1. Demographics and anthropometrics (age, sex, race, ethnicity, height, and weight);
2. Cardiovascular factors, procedures, and related drugs (e.g., diabetes, hypertension, beta-blocking agents, etc.);
3. Mental health disorders;
4. Physical trauma (e.g., head injury, fall);
5. Gastrointestinal disorders, procedures, and related drugs (e.g., colonic lesion, diverticulosis, colonoscopy, etc.);
6. Neurologic and ophthalmologic disorders;
7. Bone or muscular disorders (e.g., osteoarthritis);
8. Infection and related drugs (e.g., cholera, cough suppressants);
9. Analgesia;
10. Inflammatory disorders and related drugs; and
11. Miscellaneous (ear/nose/throat disorder, vitamin B12, flu vaccine, and mammography).

## Discussion

We achieved a high-performance random forest model with an AUC of 0.84 that is comparable with previous EHR prediction models but extends the prediction window to 10 years or more prior to diagnosis. This is the first demonstration of such performance over such a lengthy time period of EHR. Other models have achieved similar performance(14-16), and a recent study showed an AUC of 0.85 for 5 years before diagnosis(18).

The most comparable analysis was conducted by Li et al.(18), which evaluated multiple time windows from 0 to 5 years before onset and predicted AD with high accuracy (AUC 0.94 and 0.85 for 0 and 5 years, respectively). Comparing their model with ours, we achieved similar PPV (0.40 compared to their model 0.49) and NPV (0.92 compared to their model 0.93) showing that even at 10 years out, we can achieve similar predictive values. As in their study, we can pinpoint controls with a fair degree of accuracy, but it is more difficult to identify those with AD. In part, this may be due to the EHR’s capacity to accurately diagnose AD cases, where past studies have shown high accuracy but not perfect accuracy with PPV of ∼90% for EHR vs. gold standard(23). One recent study used expert guided diagnosis of AD and found similar performance when using the EHR to predict AD onset 7 years prior(19). EHR case data may help identify potentially high-risk patients for monitoring (e.g., cognitive screening), lifestyle interventions that could delay progression, and screening or confirmatory biomarker and neuroimaging studies. Much work remains prior to implementing such models in clinical settings, including improving the predictive capacity, but this is a first step in that direction. In part, these models may be improved through biomarker confirmed dementia especially AD. They may also be potentially improved by predicting more complex diagnostic categories, e.g., predicting different dementia diagnoses and other related disorders.

In general, many factors were predictive, including demographic, cardiovascular, inflammatory, and mental health as well as certain procedures. These are well in line with past models of AD that have demonstrated many of these as predictive factors(24, 25), including age, weight/BMI, height (which may be an indicator of frailty in old age(26)), head injury, cardiovascular disease (and related factors), hearing or vision impairment, untreated depression and psychosis, and sedentary lifestyle.

A recent study identified many of these factors, including features that overlapped, such as age, cerebrovascular disease, mood disorder, diabetes, ischemia, weight/BMI, race, and confusion(18). They additionally found factors we did not find, including fatigue, malaise, debility, and epilepsy (18). Using a deep phenotyping approach combined with EHR data, another group showed similar factors that predicted AD, including hypertension, hyperlipidemia, osteoporosis, and mental health disorders(27). It is important to note that some of these may be related to social determinants of health – for example, those who undergo preventative screenings are generally wealthier, educated, have insurance, and may be more likely to adopt healthy behaviors. Some screenings are also associated with age, which itself is predictive of AD. Li et al.(18) similarly identified features found using our modeling approach, including age, diabetes, mammography, blood pressure, hypertension, mood disorders, seizure, vascular disorders, confusion, sleep apnea, and race. They additionally identified routine medical exams and naproxen use, which we did not. While they looked at the top 20 most important features, we listed a larger set of features based on the feature importance. In concurrent research conducted by our team, as reported in a recent study by Malec et al. (28), sleep apnea has been recognized not only as a confounding variable that complicates the relationship between depression and AD but also as a potentially modifiable risk factor for AD. Furthermore, sleep apnea may independently contribute to the risk profile for AD.

This convergence helps identify features that may be important for predicting AD. However, prediction is not causal, and none of these models, including ours, identify causal mechanisms for any identified features. Indeed, we present the features and their PFI data simply to demonstrate the feasibility of applying machine learning tools to EHR data to identify and predict early risk; we cannot rank these features or discuss their relevance to future AD diagnosis as part of this project. In a recent study(19) that used EHR to predict AD 7 years prior to onset, they additionally conducted matched analysis and prediction and found that certain features were non-specific, e.g., hypertension as they were no longer predictive after conducting matching.

Various approaches have been suggested for increasing the time window for preventive brain health services(29, 30), such as early risk assessment through *APOE* genotyping and lifestyle factors that may contribute to risk (30). Still, a major challenge is identifying *who* should receive these risk assessments. Embedding machine learning and risk prediction models directly into the EHR could facilitate this process. The EHR could help identify high-risk patients so that they could undergo cognitive testing and eventually biomarker screening, with those showing early changes in brain health referred to specialty clinics. An embedded EHR tool could also aid in continuous passive monitoring of risks to brain health. Our results complement two recent findings that estimated the timing of various AD markers to time of onset in the US(31) and China(32) that showed peripheral and central markers of amyloid accumulate 15 to 19 years prior to diagnosis, markers of tau, (peripheral) neurodegeneration, and inflammation 10 to 14 years prior, and markers of neural atrophy 7 to 9 year prior. This is especially relevant with the advent of highly accurate plasma markers of AD.

One major challenge of many studies in AD is identifying patients who are at-risk from the community. Often cross-sectional, longitudinal, and clinical trial study staff call and screen thousands of patients before identifying participants who are interested in participating but also are good candidates. The EHR may help facilitate this process by identifying these participants. In the Lecanemab clinical trial(33), they screened 5967 participants with 1795 participants recruited. They found that 4172 participants met exclusion criteria or ∼70% of those screened. The EHR may not only identify patients who are at high risk but also increase the fidelity of this process by identifying these patients. In addition to cost, there is a significant cost associated with this recruitment process that could be reduced by identifying this high-risk population. Finally, even when considering longitudinal studies of AD, if we can identify high-risk populations automatically, we can recruit participants who are at high-risk of cognitive decline and conduct longitudinal studies to better understand mechanisms of AD. This could potentially reduce the sample size needed for many large studies but also reduce the costs associated with recruitment.

It is important to note that while certain features identified may not be immediately understandable, these data are 10 years prior to the onset of AD. Thus, features like ‘impaired cognition’ are relevant predictive factors. In addition, features like ‘psychotic disorder’ were not excluded as our goal was to predict AD diagnosis even if psychotic disorders may be a sign of existing AD. Ultimately, the EHR is not perfect, and diagnostic codes are not always accurate, but our model conducts these predictions under such assumptions.

There are several limitations of the current work. The model should be validated in independent data and be further validated clinically for utility. While the current model uses data from a single large health network that is remarkable for having a large number of patients who have remained within the same geographic area for many years, many health system EHRs have a very limited number of samples or years of continuous records, which could limit generalizability as an EHR-embedded tool. There is a need to expand the testing of this model to better assess generalizability. The EHR also does not have certain types of information embedded in it like measures of social isolation though this may be in notes, which is a limitation. We used records from 2016 onward to identify AD patients to leverage the more robust AD dementia diagnostic guidelines available in that time frame to improve case identification. In addition, EHR diagnoses are thought to fairly accurately be associated with gold-standard diagnostics(23). While the current model predicts AD, extending this to other dementias may be important for performance validation. The ICD codes used here did not correspond fully with validated EHR phenotypes, which may be a limitation. By only looking at those patients with at least 10 years of data and by using a case-control design, there may be issues related to bias of the features and samples. The primary way to address such problems is to test these models in multiple EHR systems.

In conclusion, our work is significant for showing that it is feasible to predict AD diagnosis 10 years prior to onset using EHR data. The random forest model achieved a relatively high AUC and identified factors that predicted future AD diagnosis. The AD prediction factors set included known and potentially novel factors. While work remains to be done on the refinement and generalizability of these models, we believe they can be used to help identify and monitor at-risk populations and potentially to enable early preventive or therapeutic interventions. Given the long course of AD, this work suggests it may be possible to implement lifestyle interventions to reduce long-term risk over the course of a much longer span of time (i.e., 10 years).

## Data Availability

All data produced in the present study are available upon reasonable request to the authors and with compliance with the appropriate data use agreements and protections of the data.

## ^a^Abbreviations

AUPRC: area under the precision-recall curve
CDM: Common Data Model
OHDSI: Observational Data Health Sciences and Informatics
PFI: permutation feature importance

## Data Statement

The data has not been previously presented orally or by poster at scientific meetings.

## Acknowledgments

We would like to thank Elizabeth Wu for her contribution to this manuscript.

## Funding

US National Institutes of Health grants supported this research, including T15LM007059, K99LM013367, R00LM013367, UL1TR001857, K01AG071849, and T32AG055381, as did a Pitt Momentum Fund Teaming Grant from the University of Pittsburgh Office of the Provost. The views expressed here do not necessarily reflect the views of the National Institutes of Health.

## Author Contributions

Conceptualization: RDB, SAM, SMA, CES, ASL, PM, JCS, MK, HTK. Data Curation: SBT, RDB, SAM, SMA, CES, PM, JCS, HTK. Formal Analysis: SBT, RDB, HTK. Drafting: SBT, RDB, HTK. Writing: SBT, RDB, SAM, SMA, CES, ASL, PM, JB, JX, DM, KS, JCS, MK, HTK.

## Conflicts of Interest

CES reports that she is Co-chair of the Sex and Gender Special Interest Group of the Diversity and Disparities PIA in ISTAART and a member of the ISTAART Advisory Council. All other authors report no conflicts of interest.

## Consent Statement

The University of Pittsburgh Institutional Review Board approved this study as exempt and confirming that consent was not necessary. Data were de-identified to a HIPAA-limited dataset.

## Notes

### Competing Interest Statement

The authors have declared no competing interest.

### Funding Statement

The US National Library of Medicine grants supported this research, including 5 T15 LM007059-32 and K99 LM013367-01A1 and UL1 TR001857; US National Institutes of Aging grant T32 AG055381; and the University of Pittsburgh's Momentum Funds. The views expressed here do not necessarily reflect the views of the National Library of Medicine of the National Institutes of Health.

### Author Declarations

Ethics committee/IRB of the University of Pittsburgh gave ethical approval for this work, ruling that it is not human subjects research.

### Summary of Updates

This version adds the column to Table 2 showing 100 * PFI / AUC (training) from the kneepoint analysis now described in the methods. Wording in Conflicts of Interest slightly modified. Figure 2 (Precision-Recall curve) removed.

